# Evaluating the use of dose-sparing vaccination strategies for Monkeypox

**DOI:** 10.1101/2022.11.04.22281966

**Authors:** Dobromir Dimitrov, Blythe Adamson, Laura Matrajt

## Abstract

The spring-summer 2022 monkeypox outbreak had over 50,000 cases globally, most of them in gay, bisexual, and other men who have sex with men (MSM). In response to vaccine shortages, several countries implemented dose-sparing vaccination strategies, stretching a full-dose vaccine vial in up to 5 fractional-dose vaccines. Recent studies have found mixed results regarding the effectiveness of the monkeypox vaccine, raising the question of the utility of dose-sparing strategies. We used an age- and risk-stratified mathematical model of an urban MSM population in the United States with approximately 12% high-risk MSM to evaluate potential benefits from implementing dose-sparing vaccination strategies in which a full dose is divided in 3.5 fractional-doses. We found that results strongly depend on the fractional-dose vaccine effectiveness (VE) and vaccine supply. With very limited vaccine available, enough to protect with a full-dose approximately one-third of the high-risk population, dose-sparing strategies are more beneficial provided that fractional-dose preserved at least 40% of full dose effectiveness (34% absolute VE), projecting 13% (34% VE) to 70% (68% absolute VE) fewer infections than full-dose strategies. In contrast, if vaccine supply is enough to cover the majority of the high-risk population, dose-sparing strategies can be outperformed by full-dose strategies. Scenarios in which fractional-dosing was 34% efficacious result in almost three times more infections than full-dosing. Our analysis suggests that when monkeypox vaccine supply is limited and fractional-dose vaccination retains moderate effectiveness, there are meaningful health benefits from providing a smaller dose to a larger number of people in the high-risk population.

## Introduction

The World Health Organization (WHO) declared the monkeypox outbreak a public health emergency of international concern on July 23, 2022 ^1^. By September 2022, there were over 50,000 cases globally, and over 21,000 cases in the United States (US) most of them occurring in gay, bisexual, and other men who have sex with men (MSM)^2^. In the US, two vaccines are approved for prevention of monkeypox: the JYNNEOS vaccine (MVA vaccine) and the ACAM2000 vaccine. JYNNEOS is currently preferred and utilized as it has fewer side effects and contraindications. Due to the shortage of vaccine supply, regulatory agencies including the US-Federal Drug Administration (FDA) authorized a lower-than-standard dose regimen of two intradermal injections, allowing each vaccine vial to be divided in up to 5 fractional-dose vaccines ^3 4,5^. This dose-sparing strategy was based on a clinical study done in 2015,^6^ but recent studies have found mixed results for effectiveness^7^ raising the question about whether fractional dosing is the best use of limited supply of MVA vaccine. In the present work, we use a mathematical model to explore different scenarios under which fractional dosing would be the optimal use of available MVA vaccine.

## Results

To compare the health outcomes from full-dose versus dose-sparing monkeypox vaccine strategies, we adapted a previously developed, calibrated, and published infectious disease dynamic transmission model of sexual activity patterns and virus spread in a population of men who have sex with men (MSM) in Seattle, WA^8^. The population included 65,000 men stratified into age- and risk-groups, with approximately 8,000 individuals in the group with high-risk of monkeypox acquisition and greater need for vaccination. A complete model description is included in the Supplement.

In the main scenario, we simulated vaccination campaigns over 5 weeks with either 2,500 or 7,500 full-dose vaccine vials available (31% and 94% of the high-risk population). For dose-sparing strategies, we assumed that each vaccine vial could be divided into 3.5 fractional-doses ^9^, so that there was enough vaccine to cover 8,750 or 26,250 people (corresponding to 2,500 and 7,500 vaccine vials respectively). We assumed that a full-dose of MVA vaccine had a vaccine effectiveness (VE) of 85% against monkeypox infection ^10^. We simulated a low (34% absolute VE) and high (68% absolute VE) fractional-dose VE scenarios, corresponding to retaining 40% or 80% of the effectiveness the full-dose vaccine (Table 1). We ran additional scenarios with 5,000 or 10,000 full-doses available, starting the vaccination campaigns with 5- or 10-weeks delay and assuming a fractional-dose VE ranging from 17% to 85% (Fig. 1). In all scenarios, the high-risk population was vaccinated first, and additional doses (if available) were then given to the low-risk population.

**Table 1.** Comparison of monkeypox vaccine full-dose to dose-sparing strategies for main public health scenario with results

|  | **Full-Dose**<br>**Vaccination**<br>**Strategy** | **Dose-Sparing Vaccination Strategy (%)**<br>Incremental difference compared to full-dose) |  |
| --- | --- | --- | --- |
| Population size of men who have sex with men [number high risk] |  | **65,000**<br>**[7924]** |  |
| Number of high-risk people vaccinated | **2500** | **8,750 (+250%)** |  |
| Vaccine effectiveness (VE) | **85%** | Low VE<br>**34%** | High VE<br>**68%** |
| Peak weekly number of new MPX infections over 6 months | 745 | 736 (-1.2%) | 170 (-77.1%) |
| Cumulative number of new MPX infections over 6 months | 5424 | 4743 (-12.6%) | 1679 (-69.0%) |
| Number of people vaccinated | **7,500** | **26,250 (+250%)** |  |
| Vaccine effectiveness (VE) | **85%** | Low VE<br>**34%** | High VE<br>**68%** |
| Peak weekly number of new MPX infections over 6 months | 115 | 655 (+465%) | 122 (+5.3%) |
| Cumulative number of new MPX infections over 6 months | 1420 | 4147 (+193%) | 1358 (-4.3%) |

**Figure 1.**
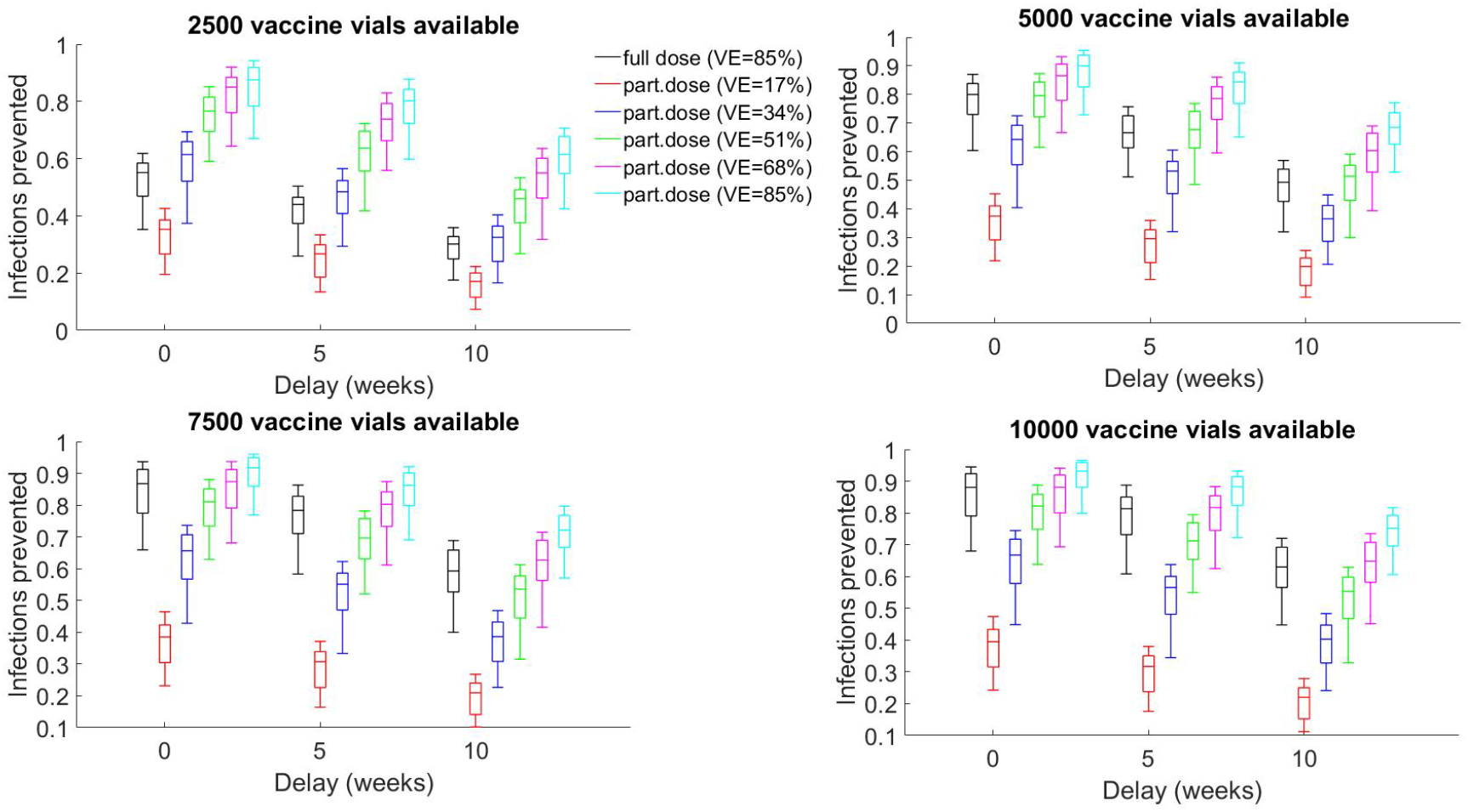
Number of infections prevented for A. 2,500, B. 5000, C, 7,500 or D. 10,000 vaccine vials available. For each panel, we compared vaccination with full-dose (VE=85%) and fractional-dose VE, ranging from 17% to 85%. We compared immediate vaccination (0-week delay) to delayed vaccination starting 5 or 10 weeks after the first infection identified.

When a limited number of vaccine vials were available (2,500 vaccine vials), sufficient to vaccinate 31% of the high-risk population, then a dose-sparing strategy prevented more infections compared to full-dose vaccination as long as the fractional-dose VE was greater than 34%, retaining 40% of the effectiveness of a full dose. In that scenario, we projected 13% fewer infections when dose-sparing strategy is implemented instead of full-dose strategy with 1% less infections at the peak recorded over 6 months (Table 1), and it averted 6% more infections than full-dose vaccination when comparing both strategies to a baseline scenario without vaccination (Fig 1A).

In contrast, if 7,500 vaccine vials were available (enough to cover 94% of the high-risk population), then full-dose campaigns were projected to outperform dose-sparing campaigns with low fractional-dose VE (34%). In that scenario dose-sparing strategies would generate almost three times more infections than full-dose campaigns, averting 21% fewer cumulative infections when compared to no-vaccination (Table 1, fig 1C).

Assuming high fractional-dose VE (68%, retaining 80% of the effectiveness of the full dose), dose-sparing strategies always outperformed or were similar to full-dose campaigns. With limited vaccine supply (2500 vials), dose-sparing strategies resulted in 69% fewer cumulative infections and 77% less infections at the peak compared to full-dose strategies, averting an additional 30% cumulative infections when compared with no-vaccination. With more vaccine supply, the number of base-case infections averted by the full-dose (86.7%) and fractional-dose (87.4%) strategies were similar with slightly more infections at the peak (+5.3%) projected with dose-sparing strategies.

In the most optimistic scenario in which fractional dosing is as effective as full dosing with limited number of vaccine vials available (2,500 vials), the fractional-dose strategy is expected to prevent more than 30% more infections (compared to no-vaccination) over 6-month period. However, in simulations with large number of vaccine vials available (at least 7500 vials), the projected increase in the infections averted by using fractional-dose compared to full-dose strategies was approximately 5% when both strategies were implemented without delay. This difference grew to 13% if vaccination was initiated after 10-week delay (Fig.1, black vs light blue). In contrast, in the more pessimistic scenario, if fractional dosing had very low VE (17%), then dose-sparing strategies would result in more infections than full-dose vaccination across all scenarios considered. The projected difference between these two strategies in proportion infection averted was projected to be between 13% to 20% with limited vaccine availability (2500 vials) but increased to more than 40% if more than 7500 vaccine vials were available (Fig.1 black vs red).

Our findings suggest that when there is limited availability of monkeypox vaccine supply there is a threshold in the fractional dose VE above which it is expected to prevent more infections than full-dose vaccination. As a greater supply of vaccine becomes available, the threshold for fractional-dose VE needed to outperform full-dose vaccination increases: with 2500 vials the threshold is below 34% as our main results showed; at 5,000 vials, fractional-dose vaccines needed to be at least 51% efficacious (approximately same proportion of cumulative e infections averted over 6 months when compared to no vaccination: 80% vs 79.5% respectively, Figure 1B). If at least 7,500 vials were available, the fractional-dose vaccines needed to be at least 68% efficacious to avert as many infections as the full-dose campaign (Fig. 1C, D). There was little gain in the number of infections averted once the available vaccine doses exceeded the size of the high-risk group (Fig.1C vs. D).

## Discussion

Several regulatory agencies have approved the use of fractional dosing for the MVA vaccine, allowing each vaccine vial of MVA vaccine to be divide into up to 5 intradermal fractional-dose vaccines. However, a recent study has raised concerns about the effectiveness of the fractional-dose vaccines^7^. In the present work, we used a previously validated model of a network of an urban MSM population in the US and adapted it to the summer of 2022 US Monkeypox outbreak. Our analysis projected better population health outcomes with a dose-sparing compared to full-dose vaccine strategy in times when there is shortage in monkeypox vaccine supply. Considering that the US has secured 1.1 million of MVA vaccines and that there are an estimated 1.7 million people at high-risk ^11^, our results suggest that dose-sparing strategies would outperform full-dose vaccination provided that the fractional-dose retains at least 40% effectiveness of the full-dose. These results are consistent with previous studies for other infectious diseases like cholera ^12 13^, influenza ^14, 15^ and COVID-19 ^16^, that have found that dose-sparing strategies are optimal when the fractional-dose VE is around half as effective as the full dosage. The fewer the doses of MVA vaccine available, the projected payoff from using dose-sparing strategies increases. Finally, we demonstrated that delaying the vaccination even by few weeks may lead to substantial reduction of the expected benefits from the vaccination campaign in terms of prevented infections. We have reported similar results when COVID vaccines were rolled out in early 2021.^17^

Our model, like any mathematical model, is subject to some limitations. We assumed that the MVA vaccine is highly efficacious in preventing infection, but more data is needed to evaluate the effectiveness of the MVA vaccine in this outbreak. If the vaccine only prevents disease but not transmission, vaccinated individuals could continue to spread the monkeypox virus to others ^2,18^. We simulated a MSM population with relatively small proportion of high-risk individuals. If this proportion was bigger, then dose-sparing strategies would be more effective when compared with full-dose vaccination for low vaccine supply, provided that the fractional dose VE remains moderately high. We assumed that monkeypox virus would transmit only in the MSM population, but spillovers to other populations are likely to happen. Reports of people getting infected without sexual exposure are starting to emerge ^19^, but continue to be rare.

Taken together, our results suggest that when monkeypox vaccine supply is limited and fractional dosing retains at least moderate effectiveness, then there can be meaningful health benefit from providing a smaller dose to a larger number of people in the high-risk population. It is then imperative to evaluate the effectiveness of fractional dose for the MVA vaccine in the context of the current outbreak.

## Data Availability

Code for this manuscript will be available at repository

https://github.com/lulelita/monkeypox

## Acknowledgments

This work was partially supported by grants from the National Institute of Allergy and Infectious Diseases of the National Institutes of Health (UM1AI068635 and UM1AI068617).

## SUPPLEMENTARY APPENDIX

### Model Description

Model of MPX transmission among sexually active MSM population (age 15-65) was developed on the basis of a model of HIV transmission previously published here.

**Figure S1.**
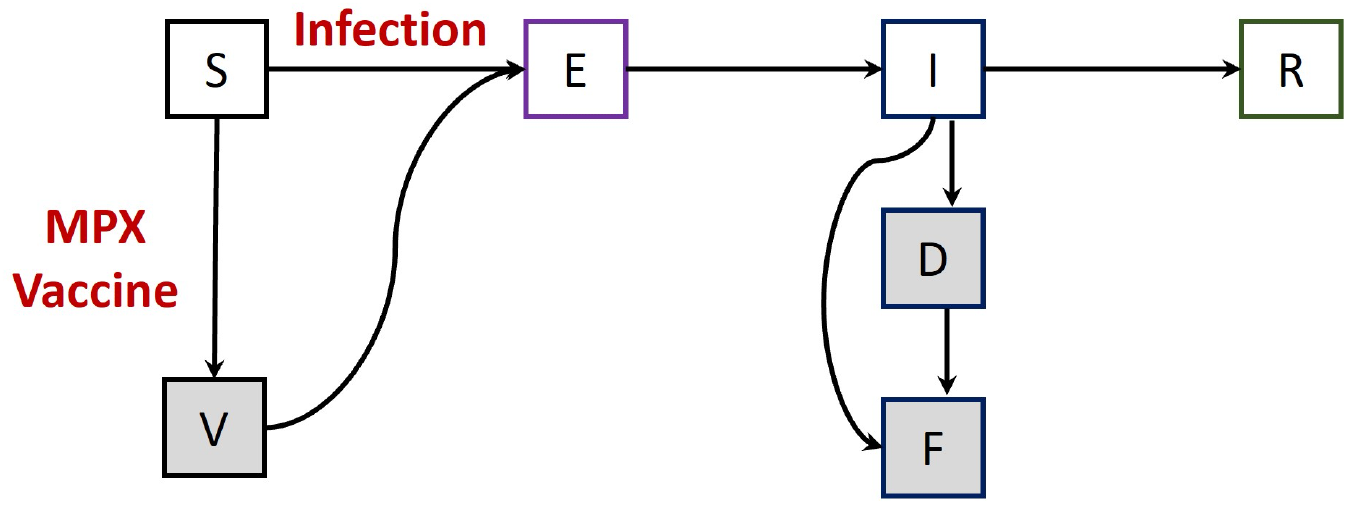
Model diagram showing MPX transmission dynamics. Population is additionally stratified by age, risk and sexual positioning.

In short, individuals are divided into risk groups by sexual activity (*i* : low risk <5 partners or high risk 5+ partners in past 12 months), age groups (*k* : young 15-24 years, middle 25-44 years, or older 45-64 years), and sexual role (*j* : insertive, receptive, or versatile position). The rate of mixing between populations depends on the current distribution of risk, role, and age groups. We ensure the overall number of partnerships between groups remains balanced by continually updating the fraction of partners someone with a particular risk, role and age group has with the rest of the population. Susceptible MSM who become sexually active join the community at constant rate, corresponding to estimated population growth among the MSM population in Seattle.

The rates at which individuals acquire MPX depends on the number of partners per susceptible person, the number of sex acts per partnership and the acquisition risk per act with infectious partner. Sexual frequency was used as surrogate measure for the rate of close contacts which facilitate MPX transmission implemented through multiplier. MSM, infected with MPX, initially move to an exposed (E) but not infectious class before becoming infectious (I) and recover (R) with complete immunity against reinfection. Infected individuals also become diagnosed at rate (σ). We assume that vaccination do not affect the course of breakthrough infections of their infectiousness.

### Model Equations

#### Parameters

In general:

- Risk status *i* ∈ {1 (low risk), 2 (high risk)}
- Role status *j* ∈ {1 (insertive), 2 (receptive), 3 (versatile)}
- Age status *k* ∈ {1 (young), 2 (middle), 3 (old)}

*d*_*k*_ : Death rate (non-HIV related) for age *k*

μ : MPX-related mortality rate

ϵ : Rate to advance from exposed to infectious class

σ : Diagnostic rate of infectious MSM

*r* : Recovery rate

*a*_*k*_ : Aging rate from age *k* to age *k* +1

*ρ*_*i*,*k*_ : Fraction of population with risk status *i* and in age group *k*

*r*_*j*_ : Fraction of population with role status *j*

*b* : Population birth (aging into population) rate

*λ*^*i, j*,*k*^ : Force of infection for newly infected population entering exposed state *E*^*i*,*j*,*k*^

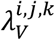 :Force of infection for newly infected vaccinated population entering exposed state *E*^*i*,*j*,*k*^

*S*^*i*,*j*,*k*^: Susceptible (unvaccinated) population with risk status *i*, role group *j* and age group *k*

*V*^*i*,*j*,*k*^: Vaccinated susceptible population with risk status *i*, role group *j* and age group *k*

*E*^*i*,*j*,*k*^: Exposed (not infectious) population with risk status *i*, role group *j* and age group *k*

*I*^*i*,*j*,*k*^: Infectious population with risk status *i*, role group *j* and age group *k*

*D*^*i*,*j*,*k*^: Diagnosed infected population with risk status *i*, role group *j* and age group *k*

*R*^*i*,*j*,*k*^: Recovered (fully protected) population with risk status *i*, role group *j* and age group *k*

*F*^*i*,*j*,*k*^: Cumulative MPX-related deaths with risk status *i*, role group

*I* : Infected population size *I* = Σ_*i*,*j*,*k*_+(*E*^*i*,*j*,*k*^ + *I*^*i*,*j*,*k*^ + *D*^*i*,*j*,*k*^).

*S* : Susceptible population size *S* = Σ_*i*,*j*,*k*_+(*S*^*i*,*j*,*k*^ + *V*^*i*,*j*,*k*^+*R*^*i*,*j*,*k*^)

*N* : Total population size *N* = *S* + *I j* and age group *k*

#### Equations

For simplicity, all variables corresponding to age group *k* = 0 have a value 0. Between vaccination campaigns:

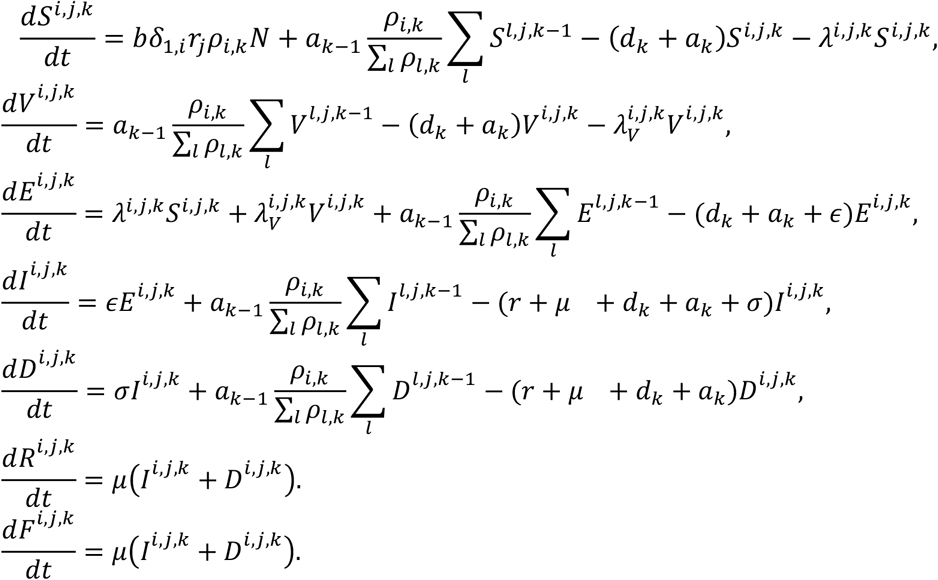

Vaccination is performed at 5 weekly campaigns in which equal number of MSM are vaccinated (Vacc_per_campaign) with high risk MSM being prioritized. The following algorithm is implemented:

1. If number of vaccinated per campaign is smaller than the unvaccinated high-risk MSM (Vacc_per_campaign <Σ_*j*,*k*_ *S*^2,*j*,*k*−1^) then only high risk MSM are vaccinated with doses distributed proportionally across age and role groups.
2. If number of vaccinated per campaign is greater than the unvaccinated high-risk MSM but (Vacc_per_campaign >Σ_*j*,*k*_ *S*^2,*j*,*k*−1^) then all high risk MSM are vaccinated with remaining doses distributed some proportionally across age and role groups of low risk MSM.

#### Force of Infection

The number of new infections among the susceptible classes X^*i*,*j*,*k*^ (risk status *i*, role status *j* and age group *k*), where X could be S or V, due to contacts with the infectious class *W*^*x*,*y*,*z*^ (risk status *x*, role status *y* and age group *z*), where W could be I or D, are calculated using the following:

|  |  |  |  |  |  |  |  |  |
| --- | --- | --- | --- | --- | --- | --- | --- | --- |
| Number<br>susceptible<br>in class $X^{i,j,k}$ | X | Estimated<br># partners<br>for age<br>group $k$<br>with risk<br>status $i$ | X | likelihood that<br>the partner is<br>from age group<br>$z$ with risk status<br>$x$ and role status<br>$y$ | X | probability that<br>the partner is<br>infected in state<br>$W$ | X | Annual<br>transmission<br>probability in<br>partnership<br>between<br>$X^{i,j,k}$ and $W^{x,y,z}$ |

Partnership likelihood is based on mixing matrices by age, risk and role, i.e, proportion of partnership that a person from each group has with every other group. These matrices are updated at each step to balance the participating parties (see below). Age/risk mixing is assumed independent from role mixing.

As a result, the force of infection on a susceptible individual who is not vaccinated is:

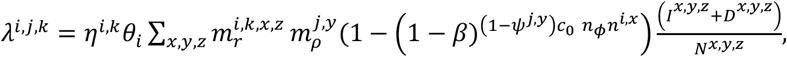

while the force of infection on a susceptible individual who is vaccinated is:

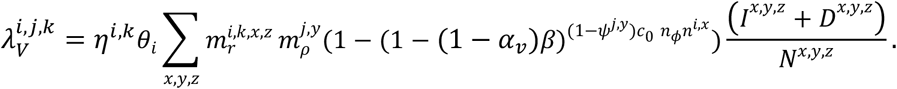

Here:

*η* ^*i*,*k*^ : number of partners for individuals in risk group *i* and age group *k*,

*n*^*i*,*x*^ : number of sexual acts per year in partnership between risk group *i* and risk group *x*,

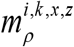 :probability for a partner of a person with risk status *i* from age group *k* to be with risk status *x* from age group *z* (risk/age mixing),

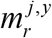 :mixing probability between people with role *j* and role *y* (role mixing),

*β* : MPX-transmission risk per contact from infectious MSM to uninfected unvaccinated MSM

*α*_*ν*_: Vaccine efficacy in reducing susceptibility per act,

*N* ^*x*,*y*,*z*^ : Population size with risk status *x*, role status *y*. and age group *z*

### Partnership Balancing

#### Balancing by age and risk

We consider mixing between all possible 6 age/risk groups. Likelihood of partnerships between 2 age/risk groups are stored in 6×6 age/risk mixing matrix ***A=a***_***ij***_ representing the likelihood that each partner of an individual from group *i* to be from group *j*. ***A*** is informed initially by data representative for the MSM population in Seattle. The matrix is constantly updated to balance the number of partnerships between different age/risk groups. The procedure aims to equilize the number of people participating at each end of the partnerships between 2 different age/risk groups (*i* and *j*) calculated as:

(Number of people in group *i*)*(Number of partners per individual)*(Likelihood *a*_*ij*_)

At each time step, the balancing procedure proceeds as follows:

1. All off-diagonal pairs *a*_*ij*_ and *a*_*ij*_ are adjusted to equalize the number of people participating at each end of the partnerships. Adjusted is the entry which leads to decrease in the sum of the off-diagonal values
2. Diagonal entries are adjusted to guarantee that each row sums up to 1.

Step 1) of the procedure guarantees that the sum of the off-diagonal entries decreases and therefore remains below 1. This makes step 2) always possible. The procedure favors sexual mixing within each age/risk group which is supported by self-reported behavioral data.

**Table S1.** Initial value of the mixing matrix used in the simulations

| Mixing likelihoods | Young, low risk | Middle, low risk | Old, low risk | Young, high risk | Middle, high risk | Old, high risk |
| --- | --- | --- | --- | --- | --- | --- |
| Young, low risk | 0.2394 | 0.0986 | 0.1925 | 0.2066 | 0.0751 | 0.1878 |
| Middle, low risk | 0.1323 | 0.254 | 0.2222 | 0.0899 | 0.127 | 0.1746 |
| Old, low risk | 0.1666 | 0 | 0.381 | 0.1111 | 0 | 0.3413 |
| Young, high risk | 0.1231 | 0.0313 | 0.0901 | 0.3217 | 0.1176 | 0.3162 |
| Middle, high risk | 0.0899 | 0.1013 | 0.0727 | 0.1396 | 0.3327 | 0.2638 |
| Old, high risk | 0.0806 | 0 | 0.1573 | 0.2258 | 0 | 0.5363 |

#### Balancing by role

Fractions of partners by role are constantly updated to balance the number of partnerships between different role groups.

We use the following likelihood matrix of partnering between sexual role groups:

**Table S2.** Sexual partnership mixing matrix

| Role group | Receptive | Insertive | Versatile |
| --- | --- | --- | --- |
| Receptive | a | b | 1-a-b |
| Insertive | x | c | 1-c-x |
| Versatile | y | z | 1-y-z |

Assuming that the overall number of partners per year is the same for each role group our balancing procedure requires that:

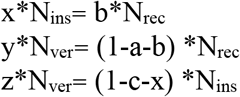

In all simulations, the values of a, b and c remain constant while x, y and z are updated at each time step. Simulations which result in negative mixing rates are discarded.

### Model Inputs

**Table S3.** Model parameters defining vaccination scenarios. included in the main analysis. **Highlighted values** were used in the scenarios listed in Table 1.

| Parameter/Description | Values |
| --- | --- |
| **Total number of full vaccine doses available** | **2,500, 5,000, 7,500** and 10,000 |
| **Number of full vaccine doses distributed weekly** | **500, 1,000, 1,500** and 2,000 |
| **Delay of the vaccination campaign** | **No delay, 5 weeks** and 10 weeks |
| **Vaccination strategy** | **Full dosing vs. fractional dosing** |
| **Vaccine efficacy in reducing the risk to acquire infection** | **85%** (full dose)<br>17%, **34%**, 51%, **68%** and 85%<br>(fractional dose) |

**Table S3.** Complete list of fixed parameter values used in the analysis.

| Variable name | Description | Value |
| --- | --- | --- |
| $N_0$ | Estimated population size in 2022 | 65000 |
|  | Initial fraction of MSM infected with MPX | 0.1% |
| $1/a_1$ | Duration of the young age group (15-24) | 10 years |
| $1/a_2$ | Duration of the middle age group (25-44) | 20 years |
| $1/a_3$ | Duration of the old age group (45-64) | 20 years |
| used in $\rho_{i,1}$ | Fraction of the population in the young age group | 0.168 |
| used in $\rho_{i,2}$ | Fraction of the population in the middle age group | 0.463 |
| used in $\rho_{i,1}$ | Fraction of MSM in the young age group with high risk status | 0.31 |
| used in $\rho_{i,2}$ | Fraction of MSM in the middle age group with high risk status | 0.099 |
| used in $\rho_{i,3}$ | Fraction of MSM in the old age group with high risk status | 0.065 |
| $d_1$ | Death rate (non- MPX related) for the young age group | 0.001319 |
| $d_2$ | Death rate (non- MPX related) for the middle age group | 0.001574 |
| $d_3$ | Death rate (non- MPX related) for the oldage group | 0.008438 |
| used in $r_1$ | Fraction of MSM with insertive role status | 0.3248 |
| used in $r_2$ | Fraction of MSM with versatile role group | 0.2683 |
| $b$ | Population recruitment rate (MSM turning 15 years) | 0.03 |
| $\sigma$ | MPX diagnostic rate on a weekly basis | 10% |
| $1/\epsilon$ | Duration of exposed (not infectious) stage | 1 week |
| $1/r$ | Duration of infectious stage ( reciprocal of the recovery rate) | 3 weeks |
| $\mu$ | Weekly MPX-related mortality rate | 1% |
| $\mu_3$ | HIV-related death rate for MSM with CD4 350-500, off ART | 0.0039 |
| $\eta^{2,1}$ | Annual number of partners for MSM in the young age group with high risk status | 10.5 |
| $\eta^{2,2}$ | Annual number of partners for MSM in the middle age group with high risk status | 10.5 |
| $\eta^{2,3}$ | Annual number of partners for MSM in the old age group with high risk status | 6 |
| $\eta^{1,1}$ | Annual number of partners for MSM in the young age group with low risk status | 1.5 |
| $\eta^{1,2}$ | Annual number of partners for MSM in the middle age group with low risk status | 1.5 |
| $\eta^{1,3}$ | Annual number of partners for MSM in the old age group with low risk status | 1 |
| $\beta$ | MPX-transmission probability per contact from infectious MSM to uninfected unvaccinated MSM | 20% |
| $m^{1,1}$ | Probability for someone in role group "insertive" to partner with someone in role group "insertive" | 0.18 |
| $m^{1,3}$ | Probability for someone in role group "receptive" to partner with someone in role group "insertive" | 0.6 |
| $m^{3,3}$ | Probability for someone in role group "receptive" to partner with someone in role group "receptive" | 0.08 |
| $\psi^{1,1}$ | Fraction of receptive acts in partnership between two MSM with insertive role status | 0.5 |
| $\psi^{1,2}$ | Fraction of receptive acts when MSM with insertive role status is partnering with MSM with receptive role status | 0 |
| $\psi^{1,3}$ | Fraction of receptive acts when MSM with insertive role status is partnering with MSM with versatile role status | 0 |
| $\psi^{2,1}$ | Fraction of receptive acts when MSM with receptive role status is partnering with MSM with insertive role status | 1 |
| $\psi^{2,2}$ | Fraction of receptive acts in partnership between two MSM with receptive role status | 0.5 |
| $\psi^{2,3}$ | Fraction of receptive acts when MSM with receptive role status is partnering with MSM with versatile role status | 1 |
| $\psi^{3,1}$ | Fraction of receptive acts when MSM with versatile role status is partnering with MSM with insertive role status | 1 |
| $\psi^{3,2}$ | Fraction of receptive acts for MSM with versatile role status partnering MSM with receptive role status | 0 |
| $\psi^{3,3}$ | Fraction of receptive acts in partnership between two MSM with versatile role status | 0.5 |
| $\alpha_v$ | MPX-vaccine efficacy in reducing susceptibility to infection when full dose is used | 85% |

**Table S4.** List of parameters and their ranges estimated in previous calibration of the HIV model in which 100 parameter sets are selected and used in this analysis.

| Variable name | Description | Range |
| --- | --- | --- |
| $\theta_1$ | Multiplicative factor which adjusts MPX contactness based on sexual activity of high-risk MSM | 10-20 |
| $\theta_2$ | Multiplicative factor which adjusts MPX contactness based on sexual activity of low-risk MSM | 15.6-20 |
| $n^{1,1}$ | Number of acts per year in partnership in which both MSM have low risk status | 40-60 |
| $n^{1,2}$ | Number of acts per year in partnership between MSM with low risk status | 2.0-5.0 |
| $n^{2,2}$ | Number of acts per year in partnership in which both MSM have high risk status | 1.0-2.0 |

## Notes

### Competing Interest Statement

The authors have declared no competing interest.

